# Immunosuppression impaired the immunogenicity of inactivated SARS-CoV-2 vaccine in non-dialysis kidney disease patients

**DOI:** 10.1101/2022.01.14.22269182

**Authors:** Yue-Miao Zhang, Xing-Zi Liu, Miao-Miao Lin, Jin-Can Zan, Yi-Tong Hu, Xiang-Qiu Wang, Wen-Qi Wu, Tai-Cheng Zhou, Hong Zhang, Ji-Cheng Lv, Li Yang, Zi-Jie Zhang

**Affiliations:** Renal Division, Department of Medicine, Peking University First Hospital, Renal Pathology Center, Institute of Nephrology, Peking University, Key Laboratory of Renal Disease, Ministry of Health of China, Key Laboratory of CKD Prevention and Treatment, Ministry of Education of China; Research Units of Diagnosis and Treatment of Immune-Mediated Kidney Diseases, Chinese Academy of Medical Sciences, Beijing, 100034, China; Center Lab and Liver Disease Research Center, the Affiliated Hospital of Yunnan University, Kunming, Yunnan 650091, China; State Key Laboratory for Conservation and Utilization of Bio-resource and School of Life Sciences, Yunnan University, Kunming, Yunnan 650091, China

## Abstract

Patients with chronic kidney disease (CKD) are at higher risk for coronavirus disease 2019 (COVID-19)-related morbidity and mortality. However, a significant portion of CKD patients showed hesitation toward vaccination in telephone survey of our center. Yet no serial data available on humoral response in patients with CKD, especially those on immunosuppression. We conducted a pilot, prospective study to survey the safety and humoral response to inactivated SARS-CoV-2 vaccine in CKD patients receiving a 2-dose immunization of inactivated SARS-CoV-2 vaccine. We found the neutralizing antibody titers in CKD patients was significantly lower than that in healthy controls, hypertension patients, and diabetes patients. Notably, immunosuppressive medication rather than eGFR levels or disease types showed effect on the reduction of immunogenicity. Interestingly, a third dose significantly boosted neutralizing antibody in CKD patients while immunosuppressants impeded the boosting effects. In conclude, our data demonstrates that CKD patients, even for those on immunosuppression treatment, can benefit from a third vaccination boost by improving their humoral immunity.

---

Patients with chronic kidney disease (CKD) are at higher risk for coronavirus disease 2019 (COVID-19)-related morbidity and mortality than general populations and, early vaccination should be prioritized for this vulnerable population. However, concerning safety and efficacy, a significant portion of CKD patients (1720/2509, 68.6%) showed hesitation toward vaccination in telephone survey of our center (**Fig S1**). Previous studies focused on exploring immune responses to COVID-19 vaccine in patients on dialysis or receiving kidney transplant.^1,2^ Yet no serial data available on humoral response in patients with CKD, especially those on immunosuppression.^3,4^

We conducted a pilot, prospective study to survey the safety and humoral response to inactivated SARS-CoV-2 vaccine in CKD patients receiving a 2-dose immunization of inactivated SARS-CoV-2 vaccine (**Item S1** and **Fig S2**). Baseline characteristics of the participants are described in **Table 1**. Briefly, the average age of CKD patients are 42.4 years with 20 (44%) females. 21 of them received SinoVac, 20 received Sinopharm and 4 received both sequentially. The most common form of CKD was chronic glomerulonephritis (27/45, 60.0%) followed by podocytopathy (5/45, 11.1%) and metabolic kidney disease (5/45, 11.1%). There were 18 patients taking immunosuppressants during the vaccination period with 17 (94.4%) receiving monotherapy. We tightly controlled age, female ratio, types of administered vaccines and sampling time across groups, except that the average age of diabetes patients is significantly older than CKD patients. None of these participants reported severe adverse effects after vaccination.

**Table 1.** Demographic and clinical characteristics of study participants. † indicates angiotensin II receptor blockers, angiotensin-converting enzyme inhibitors, insulin, metformin, acarbose, sodium-dependent glucose transporter inhibitor, calcium channel blockers, β-blocker, propylthiouracil, or levothyroxin sodium tablets.

| **Characteristics** | **Chronic kidney disease patients**<br>**(n = 45)** | **Healthy controls**<br>**(n = 100)** | **Hypertension disease controls**<br>**(n = 100)** | **Diabetes disease controls**<br>**(n = 100)** |
| --- | --- | --- | --- | --- |
| **Mean age (SD), y** | 42.4 (16.2) | 38.4 (13.3) | 39.7 (15.4) | 56.3 (8.2) |
| **Gender, n (%)** |  |  |  |  |
| Female | 20 (44.4) | 45 (45) | 47 (47) | 50 (50) |
| **Median interval between the second dose and sampling (IQR), days** | 27 (20, 33) | 30 (17, 30) | 25 (16, 38) | 28 (22, 31) |
| **Applied vaccines, n (%)** |  |  |  |  |
| SinoVac | 21 (46.7) | 48 (48) | 59 (59) | 90 (90) |
| Sinopharm | 20 (44.4) | 43 (43) | 25 (25) | 5 (5) |
| Both | 4 (8.9) | 9 (9) | 16 (16) | 5 (5) |
| **Kidney disease diagnosis, n (%)** |  |  |  |  |
| **Chronic glomerulonephritis** |  |  |  |  |
| IgA nephropathy | 21 (46.7) | - | - | - |
| IgA vasculitis | 1 (2.2) | - | - | - |
| Chronic glomerulonephritis without kidney biopsy | 5 (11.1) | - | - | - |
| **Podocytopathy** |  |  |  |  |
| Membranous nephropathy | 4 (8.9) | - | - | - |
| Focal segmental glomerular sclerosis | 1 (2.2) | - | - | - |
| **Metabolic kidney disease** |  |  |  |  |
| Diabetic nephropathy | 4 (8.9) | - | - | - |
| Hypertensive nephropathy | 1 (2.2) | - | - | - |
| **Others** |  |  |  |  |
| Uninephrectomy | 2 (4.4) | - | - | - |
| Fanconi syndrome | 1 (2.2) | - | - | - |
| Alport syndrome | 1 (2.2) | - | - | - |
| Acute interstitial nephritis | 1 (2.2) | - | - | - |
| Kidney amyloidosis | 1 (2.2) | - | - | - |
| Monoclonal gammopathy of renal significance | 1 (2.2) | - | - | - |
| Kidney stone | 1 (2.2) | - | - | - |
| **Additional diagnosis, n (%)** |  |  |  |  |
| Hypertension | 12 (26.7) | - | 100 (100) | 10 (10) |
| Diabetes | 7 (15.6) | - | 4 (4) | 100 (100) |
| Obesity | 1 (2.2) | - | - | 1 (1) |
| Gout | 3 (6.7) | - | - | - |
| Viral B hepatitis | 2 (4.4) | - | - | - |
| Fatty liver | 1 (2.2) | - | - | - |
| Coronary heart disease | 1 (2.2) | - | 2 (2) | 1 (1) |
| Asthma | 1 (2.2) | - | - | - |
| Endometrial adenomyosis | 1 (2.2) | - | - | - |
| Chronic lymphocytic leukemia | 1 (2.2) | - | - | - |
| Hypothyroidism | 1 (2.2) | - | - | 1 (1) |
| Hyperthyroidism | - | - | - | 1 (1) |
| Cerebral infarction | - | - | - | 1 (1) |
| Endometrial carcinoma of uterus | - | - | - | 1 (1) |
| **Medication exposure, n (%)** |  |  |  |  |
| Prednisone | 4 (8.9) | - | - | - |
| Bortezomib+dexamethasone | 1 (2.2) | - | - | - |
| Hydroxychloroquine | 7 (15.6) | - | - | - |
| Cyclosporin A | 3 (6.7) | - | - | - |
| Mycophenolate mofetil | 1 (2.2) | - | - | - |
| Tripterygium wilfordii | 2 (4.4) | - | - | - |
| No immunosuppression† | 27 (60) | - | 100 (100) | 96 (96) |

† indicates angiotensin II receptor blockers, angiotensin-converting enzyme inhibitors, insulin, metformin, acarbose, sodium-dependent glucose transporter inhibitor, calcium channel blockers, β-blocker, propylthiouracil, or levothyroxin sodium tablets.

Using neutralizing antibody titer of 2 as seroconversion cutoff, we found 84% (38 of 45) of CKD patients seropositive, which was lower than that in healthy controls (98%), hypertension patients (98%) and diabetes patients (95%). The median neutralizing antibody titers in CKD patients was 6.29 (IQR, 2.78-14.62), which was significantly lower than that in healthy controls [8.91 (IQR, 6.14-16.01), P = 5.15×10^−3^], hypertension patients [8.66 (IQR, 5.24-15.68), P = 0.02], and diabetes patients [9.14 (IQR, 4.62-16.22), P = 0.04] (**Fig 1A**). Conversely, we did not observe SARS-CoV-2-specific IgG or IgM differences between CKD patients and controls (**Fig 1 B-C**), despite of strong association between neutralizing antibody and SARS-CoV-2-specific IgG levels (**Fig S3**). To better understand the immunogenicity of inactivated SARS-CoV-2 vaccine in CKD patients, we further stratified CKD patients into subgroups according to their disease diagnosis, eGFR levels and medication status (receiving immunosuppressants or not). Notably, immunosuppressive medication (**Fig 1D**) rather than eGFR levels (**Fig 1E**) or disease types (**Fig S4**) showed effect on the reduction of immunogenicity. There were only 72.2% (13/18) of CKD patients receiving immunosuppressants tested seropositive after 2-dose vaccination. Moreover, we observed an immunosuppressant--dependent association between neutralizing antibody level and eGFR after adjusting for age and gender (r = 0.627, P = 0.02), suggesting that immunosuppressive agents could sensitize neutralizing antibody response to kidney function in non-dialysis kidney patients. Interestingly, a third dose significantly boosted neutralizing antibody in CKD patients while immunosuppressants impeded the boosting effects (**Table S1** and **Fig S5**).

**Fig 1.**
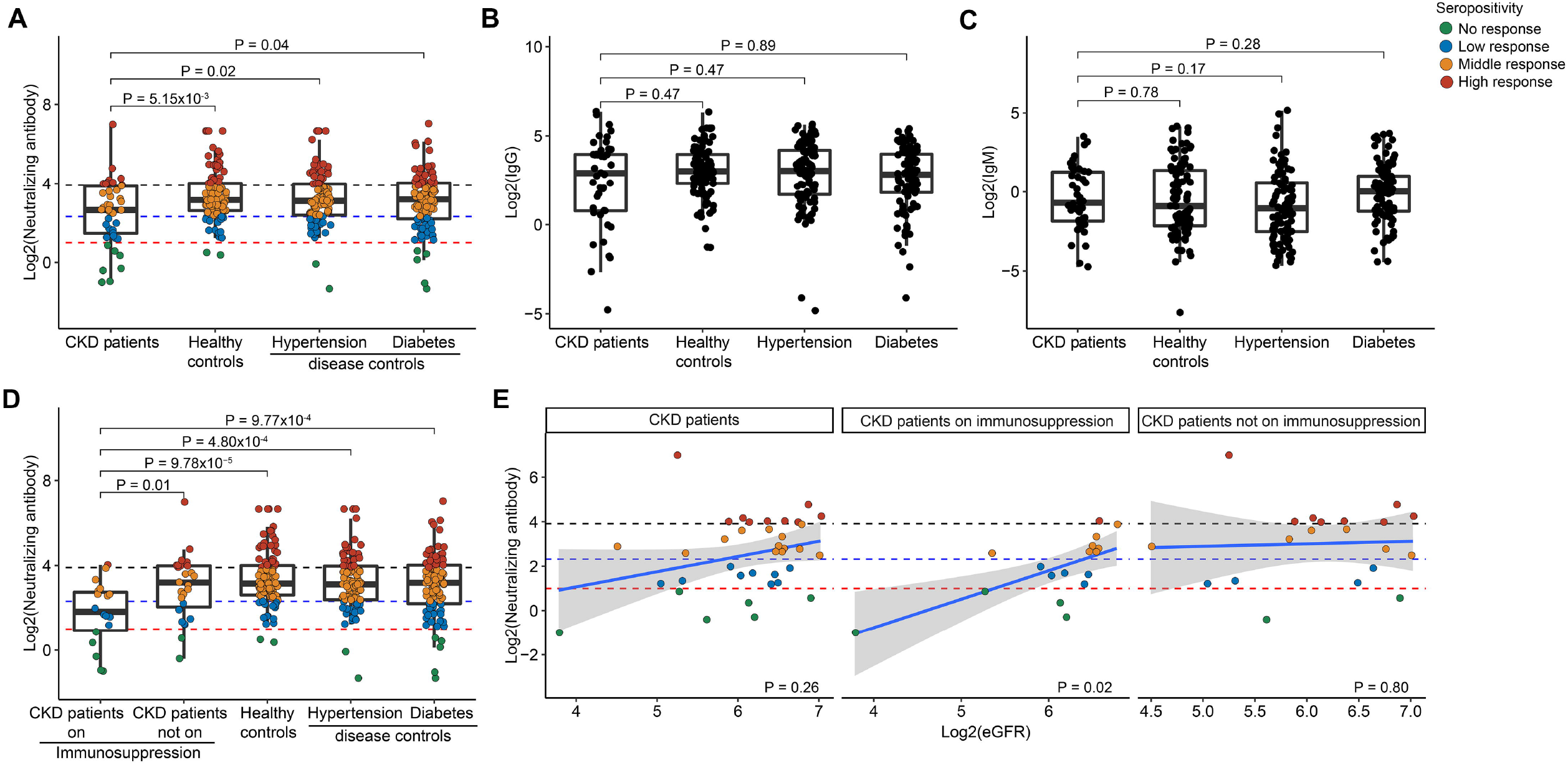
Immune responses after 2-dose inactivated SARS-CoV-2 vaccination in patients with chronic kidney disease. (**A**) Neutralization antibodies response. (**B**) SARS-CoV-2-specific IgG response. (**C**) SARS-CoV-2-specific IgM response. (**D**) Neutralization antibodies response in patients on immunosuppression. (**E**) The correlations between neutralizing antibodies and neutralization antibodies. The thresholds for neutralization antibodies is represented by the dashed lines, with <2.0 classified as no response, <5.0 as low response, <15.0 as middle response, and >=15.0 as high response. CKD: chronic kidney disease.

Our initial analysis showed that majority (84%) of CKD patients acquired detectable neutralizing antibody against SARS-CoV-2 without severe adverse effects, while the antibody titers were lower than controls. In contrast, we did not observe such difference in SARS-CoV-2-specific IgG or IgM. The deviance between neutralizing antibody and SARS-CoV-2-specific IgG responses in CKD patients indicates that neutralizing antibody rather than IgG might be the most important marker reflecting humoral immune response in CKD patients. Subgroup analyses showed that immunosuppressive therapies rather than eGFR levels or disease diagnosis impair SARS-CoV-2 vaccine-induced immunity. Our analysis in non-dialysis kidney disease patients greatly supplemented previous studies on immunosuppressive therapies^5-7^ and dialysis^8-10^ impaired vaccine responses. We found that taking immunosuppressants hampers neutralizing antibody response in CKD patients and sensitizes neutralizing antibody response to kidney function. Additionally, our data demonstrates that CKD patients, even for those on immunosuppression treatment, can benefit from a third vaccination boost by improving their humoral immunity.

## Data Availability

All data produced in the present study are available upon reasonable request to the authors.

## ACKNOWLEDGMENTS

The authors thank the study participants, and clinical staff and nurses who providing help for their participation and sampling.

## Figures and tables legends

**Fig S1 Reasons for SARS-CoV-2 vaccination hesitation in patients with kidney disease**. We screened 3637 patients with kidney disease; of these, 1128 chose not to participate, 1720 refused to vaccinate, and 744 did not qualify because they had already been vaccinated before enrollment.

**Fig S2 Study flow chart**. CKD: chronic kidney disease.

**Fig S3 The correlations between neutralizing antibodies and SARS-CoV-2-specific IgG and IgM**. (**A**) The correlations between neutralizing antibodies and SARS-CoV-2-specific IgG. (**B**) The correlations between neutralizing antibodies and SARS-CoV-2-specific IgM. CKD: chronic kidney disease.

**Fig S4 Immune responses after 2-dose inactivated SARS-CoV-2 vaccination in patients with different kidney disease diagnosis**.

**Fig S5 The effect of a third inactivated SARS-CoV-2 vaccination boost**. (**A**) Enhanced immune responses after 3-dose inactivated SARS-CoV-2 vaccination. (**B**) Enhanced immune responses after 3-dose inactivated SARS-CoV-2 vaccination in chronic kidney disease patients on immunosuppression. CKD: chronic kidney disease.

**Table S1 Demographic and clinical characteristics of participants receiving 3-dose inactivated SARS-CoV-2 vaccination**. † indicates angiotensin II receptor blockers, angiotensin-converting enzyme inhibitors, calcium channel blockers, β-blocker, and levothyroxin sodium tablets.

