## Supplementary File for "Immunosuppression impaired the immunogenicity of inactivated SARS-CoV-2 vaccine in non-dialysis kidney disease patients"

**Figure S1: Reasons for SARS-CoV-2 vaccination hesitation in patients with kidney disease.**

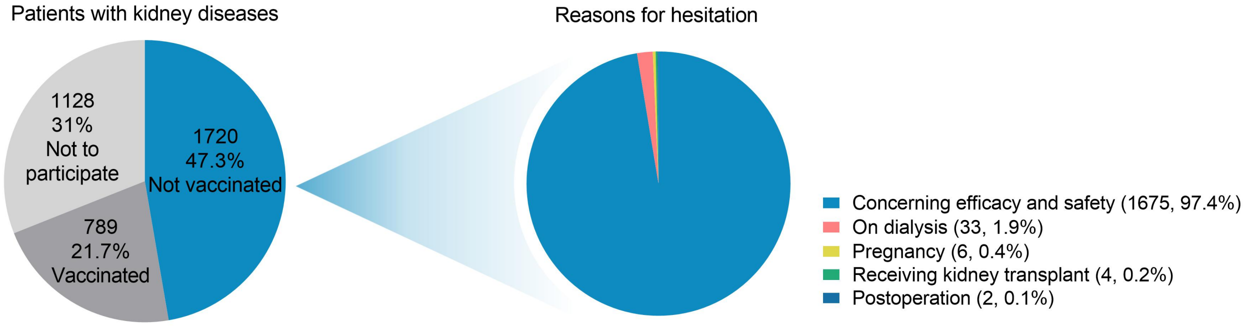


We screened 3637 patients with kidney disease; of these, 1128 chose not to participate, 1720 refused to vaccinate, and 744 did not qualify because they had already been vaccinated before enrollment.

**Figure S2: Study flow chart.**

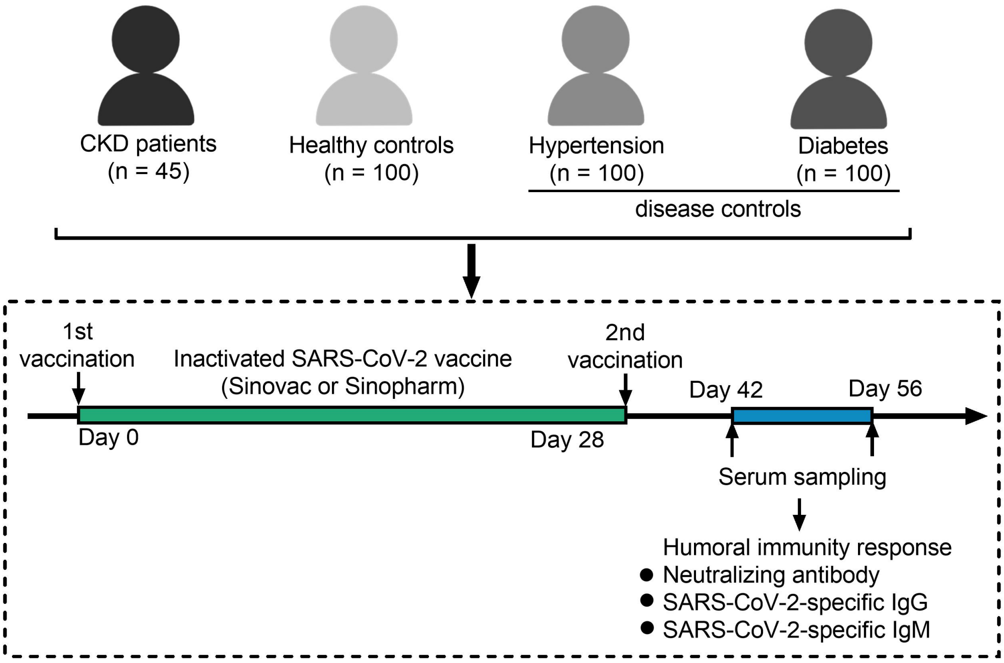


CKD: chronic kidney disease.

**Figure S3: The correlations between neutralizing antibodies and SARS-CoV-2-specific IgG and IgM.**

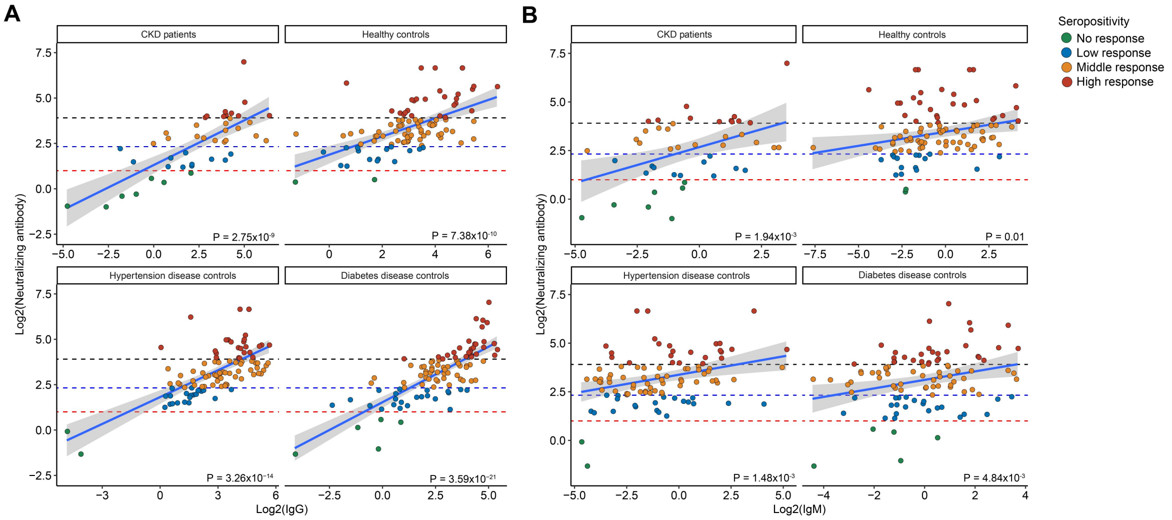


(**A**) The correlations between neutralizing antibodies and SARS-CoV-2-specific IgG. (**B**) The correlations between neutralizing antibodies and SARS-CoV-2-specific IgM. CKD: chronic kidney disease.

**Figure S4: Immune responses after 2-dose inactivated SARS-CoV-2 vaccination in patients with different kidney disease diagnosis.**

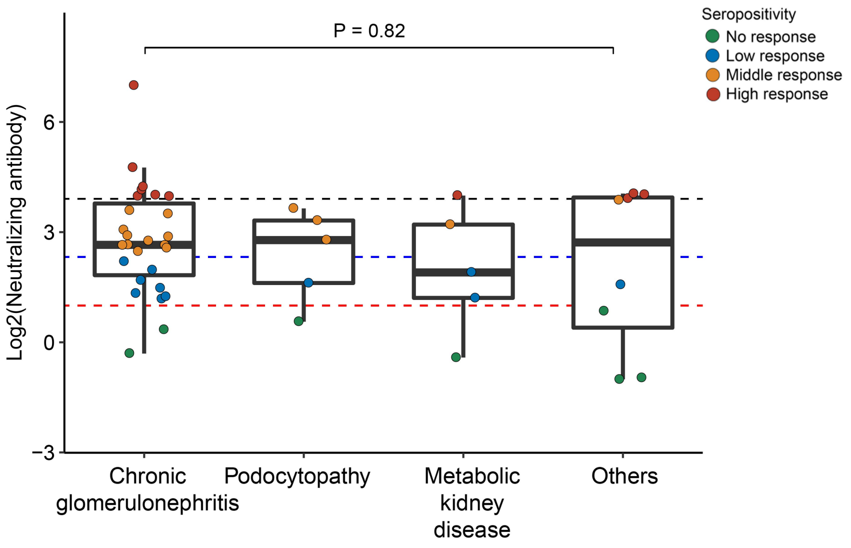

**Figure S5: The effect of a third inactivated SARS-CoV-2 vaccination boost.**

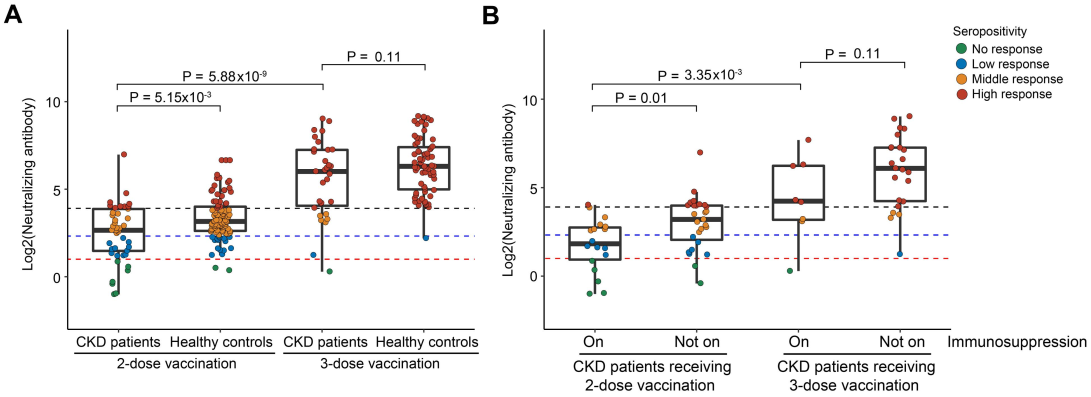


(**A**) Enhanced immune responses after 3-dose inactivated SARS-CoV-2 vaccination. (**B**) Enhanced immune responses after 3-dose inactivated SARS-CoV-2 vaccination in CKD patients on immunosuppression. CKD: chronic kidney disease.

**Item S1. Detailed methods**

*Patients:*

Between April 7 and November 1, 2021, we recruited 45 CKD patients, and 100 healthy controls, 100 hypertension patients and 100 diabetes patients with matched sampling time after vaccination at Peking University First Hospital, Yunnan University Affiliated Hospital and Center for Disease Control and Prevention in Hainan . All participants received a 2-dose immunization (3-5 weeks between two doses) of inactivated SARS-CoV-2 vaccine (SinoVac or Sinopharm). None of the participants were diagnosed with symptomatic SARS-CoV-2 infection before vaccination. Assessment of the humoral response to the vaccine was performed 20-33 days after the second dose of vaccination. We also recruited 31 CKD patients and 67 healthy controls receiving a third dose of inactivated SARS-CoV-2 vaccine to evaluate the immunological enhancement in this population. This study was approved by the Peking University First Hospital (2021[441]) and Yunnan University (CHSRE2021021).

*SARS-CoV-2 serology:*

To assess the humoral response to inactivated SARS-CoV-2 vaccines, we measured neutralizing antibody (competitive inhibition method), SARS-CoV-2-specific IgG and IgM using chemiluminescence immunoassay (MCLIA, Bioscience Co., Tianjin, China).

*Statistical methods:*

Statistics were performed using R version 3.6.1. Quantitative data were presented as mean (SD), or median (IQR: interquartile range). Qualitative data were presented as percentages. The non-parametric Wilcoxon tests were used to compare characteristics between groups.

**Table S1 Demographic and clinical characteristics of participants receiving 3-dose inactivated SARS-CoV-2 vaccination.**

| Characteristics | Chronic kidney disease patients receiving 2-dose vaccination (n = 45) | Healthy controls receiving 2-dose vaccination (n = 100) | Chronic kidney disease patients receiving 3-dose vaccination (n = 31) | Healthy controls receiving 3-dose vaccination (n = 67) |
|---|---|---|---|---|
| **Mean age (SD), y** | 42.4 (16.2) | 38.4 (13.3) | 45.4 (9.5) | 37.5 (13.1) |
| **Gender, n (%)** | | | | |
| Female | 20 (44.4) | 45 (45) | 10 (32.3) | 46 (68.7) |
| **Median interval between the second dose and sampling (IQR), days** | 27 (20, 33) | 30 (17, 30) | 18 (9, 26) | 26 (25, 32) |
| **Applied vaccines, n (%)** | | | | |
| SinoVac | 21 (46.7) | 48 (48) | 17 (54.8) | 36 (53.7) |
| Sinopharm | 20 (44.4) | 43 (43) | 14 (45.2) | 31 (46.3) |
| Both | 4 (8.9) | 9 (9) | - | - |
| **Kidney disease diagnosis, n (%)** | | | | |
| **Chronic glomerulonephritis** | | | | |
| IgA nephropathy | 21 (46.7) | - | 16 (51.6) | - |
| IgA vasculitis | 1 (2.2) | - | - | - |
| Chronic glomerulonephritis | - | - | 2 (6.5) | - |
| Chronic glomerulonephritis without kidney biopsy | 5 (11.1) | - | 7 (22.6) | - |
| **Podocytopathy** | | | | |
| Membranous nephropathy | 4 (8.9) | - | 4 (12.9) | - |
| Focal segmental glomerular sclerosis | 1 (2.2) | - | - | - |
| **Metabolic kidney disease** | | | | |
| Diabetic nephropathy | 4 (8.9) | - | - | - |
| Hypertensive nephropathy | 1 (2.2) | - | - | - |
| **Others** | | | | |
| Uninephrectomy | 2 (4.4) | - | 1 (3.2) | - |
| Fanconi syndrome | 1 (2.2) | - | - | - |
| Alport syndrome | 1 (2.2) | - | - | - |

| | | | | |
|---|---|---|---|---|
| Acute interstitial nephritis | 1 (2.2) | - | - | - |
| Kidney amyloidosis | 1 (2.2) | - | - | - |
| Monoclonal gammopathy of renal significance | 1 (2.2) | - | - | - |
| Kidney stone | 1 (2.2) | - | - | - |
| Nephrotic syndrome | - | - | 1 (3.2) | - |
| **Additional diagnosis, n (%)** | | | | |
| Hypertension | 12 (26.7) | - | - | - |
| Diabetes | 7 (15.6) | - | - | - |
| Obesity | 1 (2.2) | - | 1 (3.2) | - |
| Gout | 3 (6.7) | - | - | - |
| Viral B hepatitis | 2 (4.4) | - | - | - |
| Fatty liver | 1 (2.2) | - | - | - |
| Coronary heart disease | 1 (2.2) | - | - | - |
| Asthma | 1 (2.2) | - | - | - |
| Endometrial adenomyosis | 1 (2.2) | - | - | - |
| Chronic lymphocytic leukemia | 1 (2.2) | - | - | - |
| Hypothyroidism | 1 (2.2) | - | - | - |
| Prostate cancer | - | - | 1 (3.2) | - |
| **Medication exposure, n (%)** | | | | |
| Prednisone | 4 (8.9) | - | - | - |
| Bortezomib+dexamethasone | 1 (2.2) | - | - | - |
| Hydroxychloroquine | 7 (15.6) | - | 4 (12.9) | - |
| Cyclosporin A | 3 (6.7) | - | - | - |
| Mycophenolate mofetil | 1 (2.2) | - | - | - |
| Tripterygium wilfordii | 2 (4.4) | - | 1 (3.2) | - |
| Tacrolimus | - | - | 1 (3.2) | - |
| Tripterygium wilfordii+tacrolimus | - | - | 1 (3.2) | - |
| Paclitaxel+bevacizumab+tislelizumab | - | - | 1 (3.2) | |
| No immunosuppression† | 27 (60) | - | 23 (74.3) | - |

† indicates angiotensin II receptor blockers, angiotensin-converting enzyme inhibitors, calcium channel blockers, β-blocker, and levothyroxin sodium tablets.
